# Clinical Validation of the PSCOPE Hybrid Model Prediction of Left Ventricular Assist Device Implantation Hemodynamics: Three Patient-Specific Cases

**DOI:** 10.1101/2025.03.10.25323688

**Authors:** E. Umo Abraham, A. Welch Brett, Arman Kilic, O. Kung Ethan

## Abstract

**Objective:** The Physiology Simulation Coupled Experiment (PSCOPE) is a hybrid modeling framework designed for mechanistic cardiovascular predictive modeling. It couples a physical fluid experiment with a lumped parameter network simulation to replicate the closed-loop feedback between simulated cardiovascular physiology and fluid dynamics in the physical experiment. This study validates PSCOPE’s predictions of post-surgical physiology against clinical data in the context of HeartMate 3 left ventricular assist device implantation.

**Methods:** We designed a protocol to characterize the pre- and post-surgical hemodynamics of three adult HeartMate 3 patients using perioperative clinical measurements acquired from routine intensive care unit monitoring. For each patient, we tuned a lumped parameter network model to match their pre-surgical hemodynamic values, creating a patient-specific simulation of the pre-surgical physiology. The PSCOPE framework then modeled LVAD implantation by coupling these simulations to a physical HeartMate 3 device flow experiment. This hybrid model estimates physiological flow rate and pressure parameters to predict the patients’ post-surgical hemodynamics.

**Results:** The percentage difference between PSCOPE predictions and clinical post-surgical hemodynamics ranged from 0.0% to 44.7% across different hemodynamic parameters in different patients. The predicted cardiac index, mean pulmonary arterial pressure, central venous pressure, and pulmonary arterial wedge pressure together accurately indicated the absence of post-implant right ventricular failure in all patients.

**Conclusion:** This validation study demonstrates the potential of PSCOPE in assisting LVAD patient management. PSCOPE hemodynamic predictions could help clinicians anticipate and manage post-implant outcomes, such as right ventricular failure, thereby improving the efficacy of surgical planning.

## 1. Introduction

Cardiovascular mechanistic modeling incorporates mechanical (fluid or structural) characteristics via mathematical or physical representations in computational or experimental models, respectively. Numerical methods such as lumped parameter network (LPN), computational fluid dynamics (CFD), and finite element analysis, can be integrated into multiscale frameworks to simulate a wide range of cardiovascular phenomena including vascular pathophysiology, cardiac mechanics, and hemodynamics [1–6]. In-vitro cardiovascular modeling has advanced towards replicating various mechanistic pathways across cellular, tissue, and organ scales. Notably, 3D cell culture models provide insights into cellular responses to various stimuli, while organ-on-a-chip devices enable the study of organ-specific tissue dynamics under physiological conditions [7–9]. These methodologies recreate the microenvironment of disease-state physiology, facilitating drug discovery and development. Experimental mock circulatory loops emulate in-vivo hemodynamics under various physiological contexts, supporting direct hemodynamic investigation of anatomical phantoms and cardiovascular implants such as 3D printed vascular models, prosthetic valves and mechanical circulatory support devices [10–14]. In these setups, the physiologic context is typically specified via fixed inflow prescriptions and fluid impedances in the circuit. Hybrid experimental-computational frameworks fully integrate numerical and in-vitro models to capture the closed-loop dynamic responses between systemic-scale cardiovascular physiology and experimentally replicated mechanics [15–17]. These frameworks are especially suited for integrating simulated cardiac function with experimental fluid dynamics to attain realistic representations of hemodynamic feedback.

The translation of mechanistic cardiovascular models to clinical practice requires rigorous validation to demonstrate their accuracy and relevance in real-world scenarios. Despite rapid advancements in these modeling techniques, there is a notable lack of emphasis on validating their clinical applicability. Previous validation efforts for computational models have focused on comparisons against benchtop experimental data [18–22], rather than assessing their accuracy against in-vivo clinical data. The evaluations of in-vitro models have mostly centered on verifying their capacity to reproduce physiologically realistic parameters, but without direct clinical validation against patient data. For example, 3D cell cultures and organ-on-a-chip devices undergo histological verification to ensure microscale physiological integrity of their cellular composition and morphology [23–24], while mock circulatory loops are assessed based on their ability to achieve hemodynamic estimates within reported literature ranges for their intended physiological context [25–26]. There remains a need for in-vivo clinical validation of these mechanistic cardiovascular models to test their efficacy in guiding personalized precision care.

The Physiology Simulation Coupled Experiment (PSCOPE) is a hybrid modeling framework capable of being implemented as a mechanistic clinical predictive modeling tool. It couples an in-vitro flow circuit with an LPN simulation to model the closed-loop feedback between simulated cardiovascular physiology and fluid dynamics in a physical experiment [27–29]. Our previous study verified the PSCOPE framework by demonstrating its close agreement with a multiscale computational standard in modeling a Fontan graft obstruction in the cavopulmonary pathway [29]. While these results underscore the fidelity of PSCOPE models compared to other modeling techniques, they fall short of providing insights into its real-world clinical applicability. This study bridges that gap by clinically validating PSCOPE predictions of HeartMate 3 left ventricular assist device (LVAD) mediated physiology against patient data. These predictions could help clinicians anticipate and manage post-implant outcomes, such as right ventricular failure (RVF), demonstrating the potential of PSCOPE-guided treatment optimization.

## 2. Methods

We employ a bi-ventricular LPN to simulate the pre-surgical physiology of three adult heart failure patients. By fine-tuning the LPN’s input parameters, we synchronize its hemodynamic outputs with pre-surgical clinical measurements to generate three patient-specific LPN models known as “digital twins.” Next, the PSCOPE framework models the LVAD implantation by coupling these digital twins to a physical HeartMate 3 device flow experiment (Fig. 1). The resulting hybrid model estimates flow rate and pressure parameters characterizing the post-surgical physiology of the digital twin.

**Fig. 1.**
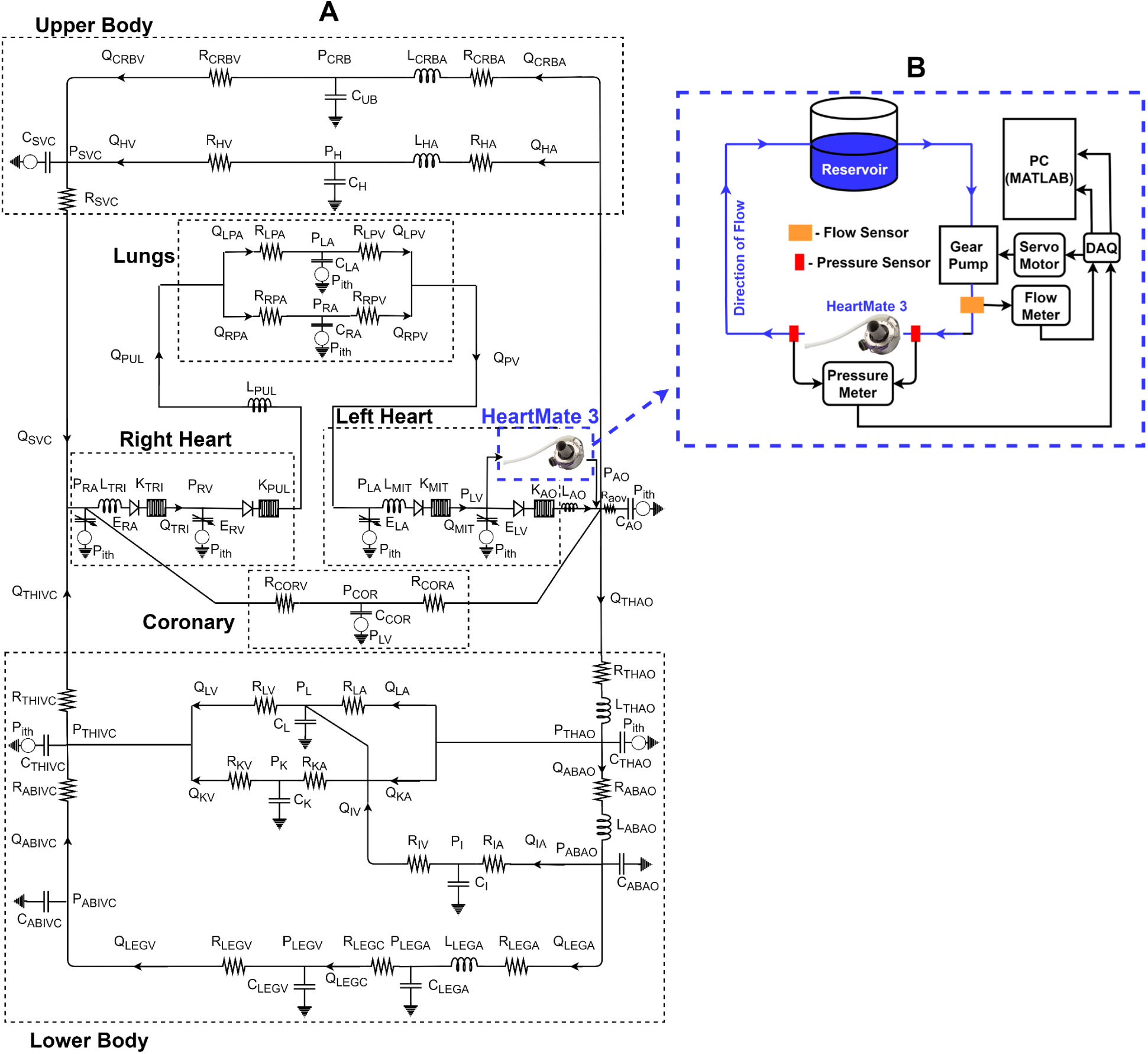
PSCOPE hybrid model of a digital twin (A) implanted with a physical Heartmate 3 LVAD (B). Digital twins simulate patient-specific preoperative hemodynamics. The HeartMate 3 is operated in an in-vitro flow loop and coupled to the digital twin.

### 2.1 Patient Cohort

The cohort consists of three adult heart failure patients who underwent HeartMate 3 LVAD implantation in 2021. The group includes two females and one male, with body surface area ranging from 1.59 m² to 2.06 m² and ages ranging from 39 to 65 years. Clinical data for each patient was sourced from the Interagency Registry for Mechanically Assisted Circulatory Support (INTERMACS) database, encompassing high-throughput in-vivo measurements acquired over several days of perioperative physiology monitoring in the intensive care unit (ICU). This data provides detailed insights into each patient’s pre- and post-surgical cardiovascular status, capturing the physiological response to the HeartMate 3 implantation.

The study was approved by the institutional review board of the Medical University of South Carolina, and informed consent was waived for each patient due to the retrospective nature of the study protocol.

### 2.2 Clinical data Characterization

The ICU health records comprise a comprehensive array of physiological parameter measurements sampled at various intervals over several days spanning the pre- and post-surgical periods. This time-series data can include thousands of data points per parameter, necessitating the derivation of characteristic values to concisely describe the patient’s pre- and post-surgical physiology. We concentrate on measurements acquired within the closest 12-hour windows preceding and following surgery. The values acquired in these windows can exhibit significant variability due to noise and physiological fluctuations surrounding the surgical event. To address noise, we identified and removed outliers in each parameter’s dataset via equation (1). Next, we characterize a representative hemodynamic state of the patient by computing the median value of each parameter within each 12-hour window (Tables 1-4). We refer to these values as the characteristic pre- and post-operative hemodynamics.

**Table 1.** Characteristic preoperative hemodynamic values derived from clinical data. HR-Heart rate; CI-Cardiac Index; LVEF-Left ventricular ejection fraction; CVP- Central venous pressure; Pao- Aortic pressure; PAWP- Pulmonary arterial wedge pressure; PAP- Pulmonary arterial pressure. Pao and PAP values are reported in “systolic/diastolic (mean)” format. “*” indicates values that were imputed due to missing clinical data.

| Parameter | Patient 1 | Patient 2 | Patient 3 |
| --- | --- | --- | --- |
| HR (bpm) | 75.0 | 84.0 | 93.0 |
| CI ($\text{l} \cdot \text{min}^{-1} \cdot \text{m}^{-2}$) | 1.9 | 2.0 | 2.7 |
| LVEF (%) | 39.0 | 20.0 | 14.0 |
| CVP (mmHg) | 10.0 | 11.0 | 6.0 |
| Pao (mmHg) | 94.0/53.0 (68.0) | 110.0/50.0 (76.0) | 104.5/69.0 (80.5) |
| PAWP (mmHg) | 25.0 | 22.0* | 21.0 |
| PAP (mmHg) | 56.0/24.0 (38.0) | 59.0/23.0 (33.0) | 55.0/22.0 (33.0)] |

**Table 2.** Comparing predicted and characteristic postoperative hemodynamic values for patient 1. Two sets of PSCOPE predictions were obtained by implementing native and adapted digital twins within the hybrid model. The percentage difference between PSCOPE predictions and clinical measurements are included.

| Parameter | Clinical | PSCOPE (native)<br>[% difference] | PSCOPE (adapted)<br>[% difference] |
| --- | --- | --- | --- |
| CI ($\text{l} \cdot \text{min}^{-1} \cdot \text{m}^{-2}$) | 1.99 | 2.37 [17.7%] | 2.44 [20.3%] |
| CVP (mmHg) | 11.0 | 9.2 [17.4%] | 8.1 [30.5%] |
| Pao (mmHg) | 83.0/66.0 (77.0) | 90.3/61.2 (82.8)<br>[8.4%/7.5% (7.2%)] | 102.1/63.8 (89.2)<br>[20.6%/3.3% (14.7%)] |
| PAWP (mmHg) | 18.0 | 11.4 [44.7%] | 16.4 [9.4%] |
| PAP (mmHg) | 34.0/18.0 (24.0) | 47.2/13.4 (27.9)<br>[32.4%/29.4% (14.9%)] | 40.0/10.8 (23.7)<br>[16.2%/50.1% (1.1%)] |

**Table 3.** Comparing predicted and characteristic postoperative hemodynamic values for patient 1. Two sets of PSCOPE predictions were obtained by implementing native and adapted digital twins within the hybrid model. The percentage difference between PSCOPE predictions and clinical measurements are included. “*” indicates values that were derived due to missing clinical data.

| Parameter | Clinical | PSCOPE (native)<br>[% difference] | PSCOPE (adapted)<br>[% difference] |
| --- | --- | --- | --- |
| CI ($\text{l} \cdot \text{min}^{-1} \cdot \text{m}^{-2}$) | 2.36 | 2.36 [0.0%] | 2.54 [7.5%] |
| CVP (mmHg) | 9.0 | 12.6 [33.1%] | 13.8 [42.2%] |
| Pao (mmHg) | 82.0/65.0 (74.0) | 111.0/51.8 (90.9)<br>[30.1%/22.6% (20.6%)] | 101.9/55.9 (84.1)<br>[21.6%/15.1% (12.7%)] |
| PAWP (mmHg) | 19.0* | 19.7 [3.7%] | 18.4 [3.1%] |
| PAP (mmHg) | 54.0/22.0 (33.0) | 62.7/20.6 (33.0)<br>[15.0%/6.7% (0.1%)] | 66.5/19.7 (33.5)<br>[20.8%/11.2% (1.5%)] |

**Table 4.** Comparing predicted and characteristic postoperative hemodynamic values for patient 1. The adapted digital twin could not be generated due to insufficient clinical data. The percentage difference between PSCOPE predictions and clinical counterparts are shown.

| Parameter | Clinical | PSCOPE (native)<br>[% difference] |
| --- | --- | --- |
| CI ($\text{l} \cdot \text{min}^{-1} \cdot \text{m}^{-2}$) | 2.33 | 2.74 [16.3%] |
| CVP (mmHg) | 9.0 | 5.9 [40.8%] |
| Pao (mmHg) | 94.0/78.0 (86.0) | 94.7/67.2 (82.1)<br>[0.8%/14.9% (4.7%)] |
| PAWP (mmHg) | N/A | 18.7 [N/A] |
| PAP (mmHg) | 35.0/17.0 (23.0) | 52.7/19.5 (30.9)<br>[40.4%/13.9% (29.4%)] |

Pulmonary arterial wedge pressure (PAWP) measurements were unavailable within the specified 12-hour windows for patient 2 (Tables 1 and 3). To address this, we impute the PAWP values via equation (2) while assuming an unchanging pressure drop “dPpul” (defined in equation (2)) between the pre- and post-operative physiology. The characteristic pre- and post-operative dPpul values were computed based on the median of mPAP and PAWP (as per equation 2) within the closest pre- and post-surgical 12hr windows (relative to the surgery), respectively, where data was available.

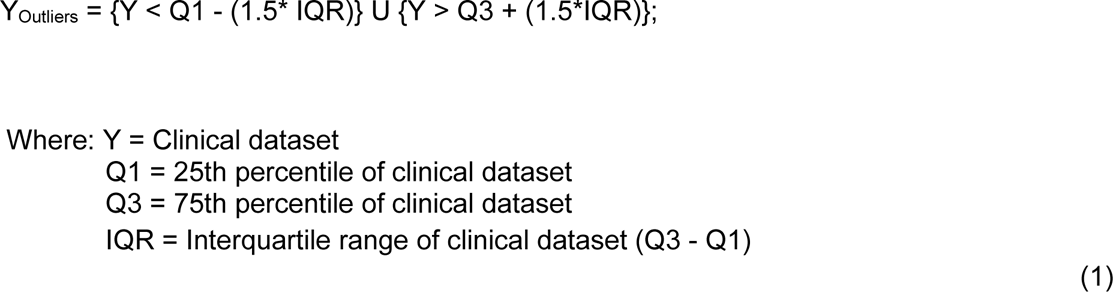

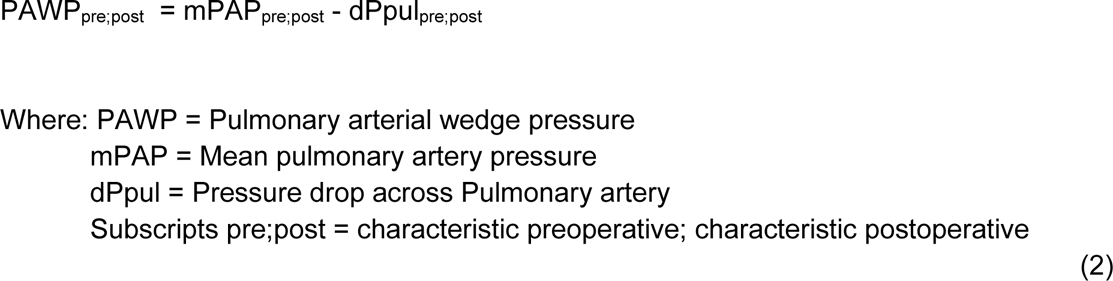

### 2.3 Digital twin: Patient-Specific LPN

A digital twin is a patient-specific LPN that simulates the patient’s reference hemodynamics. The LPN consists of cardiac and vascular circuit elements tuned to simulate a target hemodynamic profile. We constructed a generic bi-ventricular LPN based on our previous works [27;30-31] and adjusted the cardiac period and vascular resistance input values to match characteristic preoperative values for heart rate (HR), systemic vascular resistance (SVR), and pulmonary vascular resistance (PVR). The remaining input parameters were systematically tuned as per our previous work [32–33]. We generated the three digital twins in the cohort by iteratively tuning these LPN parameters until the difference between simulated and characteristic preoperative hemodynamics (Table 1) was less than 1% for each patient.

### 2.4 Virtual Surgery: PSCOPE framework

The PSCOPE framework enables the operation of a HeartMate 3 LVAD in-vitro experiment within a digital twin context by employing a coupling algorithm to facilitate feedback communication between both domains [27–29]. The in-vitro setup uses a 40% glycerin solution that mimics the density and viscosity of blood and a programmable gear pump [34; Fig. 1b] to prescribe flow rate conditions to the LVAD. The HeartMate 3 operates at the reported baseline speed for each patient case (4800 rpm - 5100 rpm) and exhibits an artificial pulse cycle that automatically pulsates flow every two seconds when the baseline speed exceeds 3900 rpm. Each pulse is characterized by the rotor speed decreasing by 2000 rpm for 0.15 seconds, then increasing by 4000 rpm for 0.2 seconds, and finally returning to baseline [35–36]. In the post-implant physiology, the interference between the artificial pulse and native cardiac cycles forms the mechanobiological cycle of HeartMate 3 mediated cardiac function. The period of this mechanobiological cycle, which we define as the coupling period, is identified in the PSCOPE framework as the shortest time interval simultaneously capturing integer multiples of simulated cardiac (digital twin), and in-vitro artificial pulse (HeartMate 3) cycles. Consequently, the hemodynamics of the PSCOPE models are analyzed over this coupling period to derive the predicted systolic, diastolic, and mean values.

We applied the coupling protocol described in our previous study [27] to identify the coupling solution for each PSCOPE model. In-vitro flow rates were monitored by scaling analog voltage outputs from a flow module (TS410 transonic system) connected to a flow sensor (16PXL transonic systems). The pressure head of the HeartMate 3 was measured via a pressure transducer (PCU 2000, Millar Instruments) connected to pressure sensors (Model DT-XX, Argon Medical Devices) adjacent to the inlet and outlet of the LVAD. We acquired all experimental flow rate and pressure voltage outputs via a NI USB-6002 data acquisition device and applied a Fourier filter to attenuate frequency components outside the frequencies of the generated flow rate boundary condition.

In each iteration of the coupling protocol, boundary conditions (periodic pressure/flow rate values) need to be prescribed over the coupling period with fixed phase-offsets relative to the HeartMate 3’s artificial pulse and digital twin’s cardiac cycles. These phase offsets determine the distinct synchronization of the Heartmate 3’s in-vitro operation with the cardiac function of the digital twin. Maintaining this synchronization between PSCOPE iterations is crucial for achieving convergence in the coupling protocol. The HeartMate 3’s artificial pulse is captured at the beginning of each iteration by recording its inlet pressure response to a reference steady flow rate boundary condition. The phase of this pulsatile pressure waveform configures the temporal operation of the programmable gear pump to prescribe periodic flow rate boundary conditions with a constant phase-offset relative to the artificial pulse cycle. The resulting periodic pressure head of the HeartMate 3 is prescribed as a boundary condition in the digital twin with a fixed phase offset relative to the simulated cardiac cycle. This ensures that the mechanobiological cycle of each PSCOPE iteration is simulated with a consistent phase. In practice, experimental noise limits the precision of the iterative phase-offset between prescribed flow rate boundary conditions and the HeartMate 3’s artificial pulse, hindering steady convergence in the coupling operation. Consequently, we specified a convergence threshold of 20% NRMSE for each PSCOPE model to balance protocol runtime and coupling solution accuracy. Once convergence is achieved, we analyze the simulated hemodynamics of the PSCOPE model to extract the predicted post-surgical physiology. The percentage difference between the predicted and the characteristic post-operative clinical data quantifies the predictive accuracy of PSCOPE for each patient. This set of percentage differences is referred to as the validation residuals of the PSCOPE model.

### 2.5 Impact of perioperative adaptations on PSCOPE predictions

The default PSCOPE method assumes unchanging postoperative LPN input parameter values compared to the preoperative values. However, LVAD implantation often induces physiological changes that can alter these parameters [37–38]; these discrepancies could adversely impact the predictive accuracy of the PSCOPE framework. To assess the effect of these perioperative adaptations on predicted hemodynamics, we compared the validation residuals of the default PSCOPE method with those obtained using updated HR, SVR, and PVR according to their characteristic postoperative values (Tables 2 and 3). The default and modified digital twins are referred to as “native’’ and “adapted’’ digital twins, respectively. This analysis focused on perioperative adaptations in HR, SVR, and PVR as these were the only LPN input parameters that can be directly computed from the clinical data. We hypothesize that incorporating these clinically observed postoperative changes into the PSCOPE models would enhance the accuracy of its hemodynamic predictions. This analysis was conducted for the two patients with corresponding clinical data (Tables 2 and 3).

## 3 Results

### 3.1 Predictions of Post-LVAD RVF

According to the latest INTERMACS guidelines, post-LVAD RVF is characterized by central venous pressure (CVP) > 16 mmHg, either measured directly or indicated by various physiological manifestations [39]. Other hemodynamic indicators, including lowered cardiac index (CI), lowered pulmonary arterial pulsatility index, and increased CVP/PAWP ratio, are also considered potentially significant markers of post-LVAD RVF [39–42]. In our PSCOPE models, we adopted the diagnostic RVF criteria characterized by CVP > 16 mmHg and CI < 2.2 L/min/m² occurring concurrently. Utilizing these criteria, the PSCOPE models accurately predicted the absence of post-LVAD RVF for all three patients in this study.

### 3.2 Prediction of Postoperative Hemodynamics

The validation residuals for mean, systolic, and diastolic values across all cases ranged from 0.0% - 44.7%, indicating variability in PSCOPE’s predictive capabilities (Tables 2-4). PSCOPE accurately predicted an increase in CI for two patients but failed to capture a counterintuitive decrease for patient 3; the validation residuals for CI in all scenarios ranged from 0.0% to 20.3%. In all cases, PSCOPE estimated a post-surgical rise in mean aortic pressure (mPao), with validation residuals ranging from 4.7% to 20.6%, and accurately replicated clinically observed trends —whether increasing, decreasing, or remaining constant— for mPAP and PAWP. The absolute differences between the predicted and characteristic postoperative values for mPAP (0.0 mmHg - 7.9 mmHg), PAWP (0.7 mmHg - 6.6 mmHg), and CVP (1.8 mmHg - 4.8 mmHg) illustrates PSCOPE’s predictive performance for key hemodynamic indicators. Lastly, the reduced accuracy in predicting hemodynamic pulsatility is reflected by the validation residuals for aortic (52.5% - 110.8%) and pulmonary arterial (27.4% - 71.4%) pulse pressures (Fig. 2B).

**Fig. 2.**
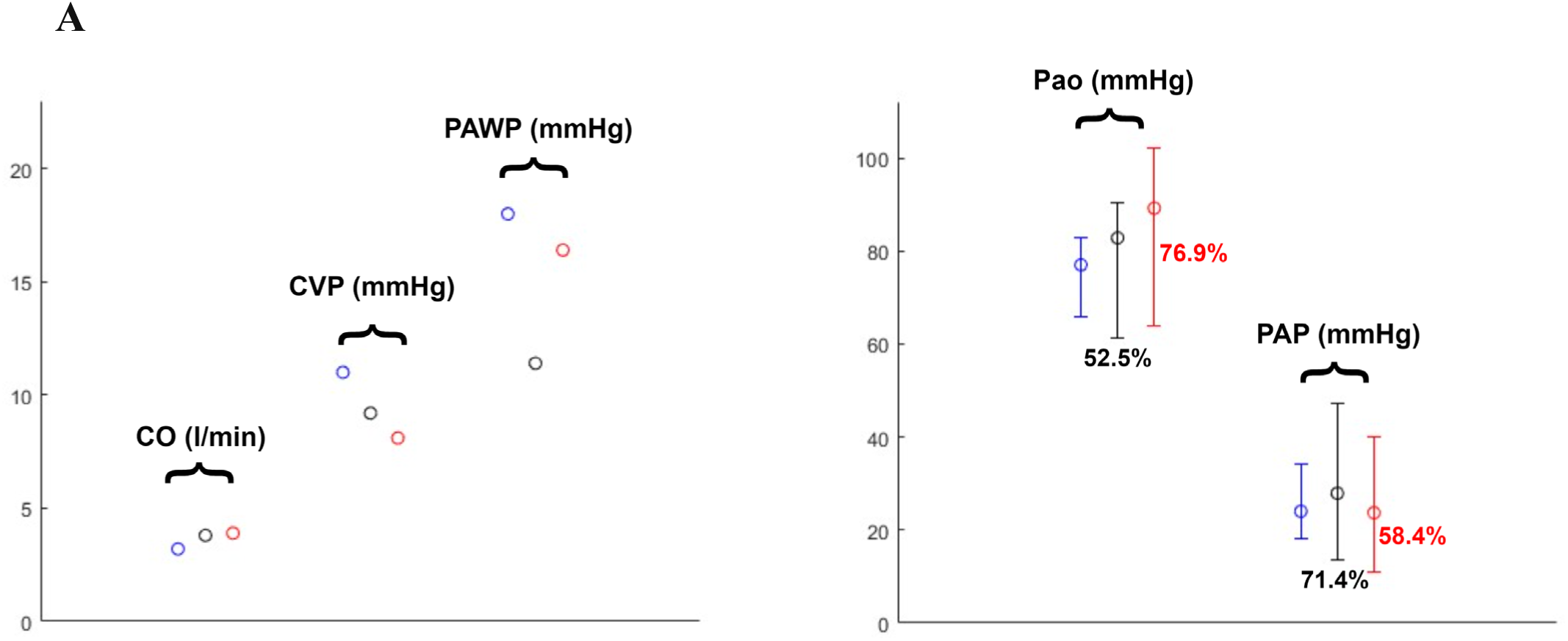
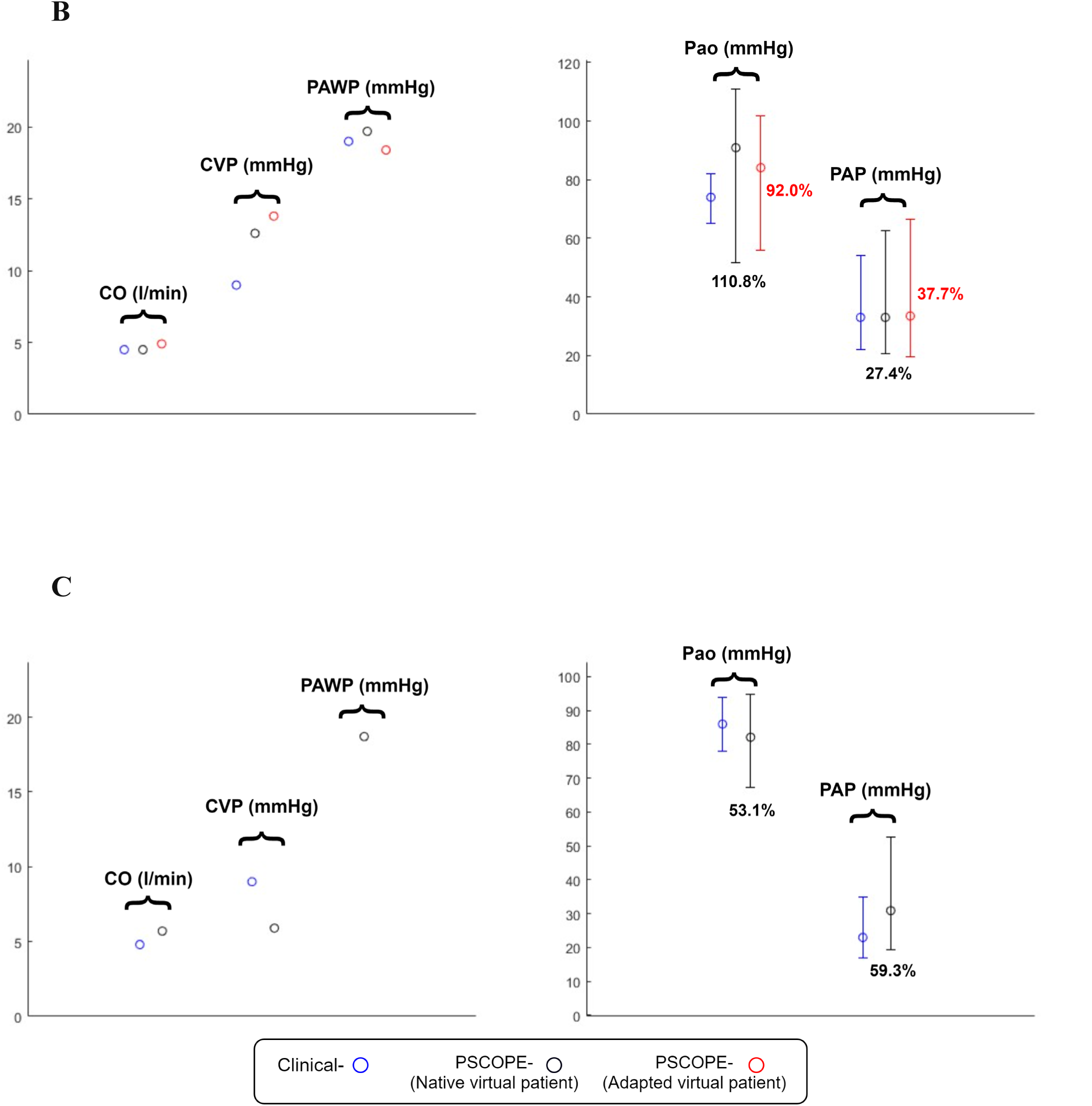
Comparing characteristic postoperative hemodynamic values to PSCOPE predictions for Patient 1 (A), Patient 2 (B), and Patient 3 (C). The percentage difference between predicted and characteristic postoperative pulse pressure is shown for Pao and PAP (B).

### 3.3 PSCOPE Model Execution and Convergence

The coupling protocol achieved the 20% NRMSE convergence threshold within 49-127 iterations across all PSCOPE models. The runtime of each PSCOPE iteration ranged from 2 to 5 mins.

### 3.4 Native vs Adapted digital twins

Comparing the hemodynamics between PSCOPE implementations of native and adapted digital twins (Tables 2 and 3) does not reveal a consistent pattern of improvement in predictive accuracy in the adapted cases. The validation residuals in the adapted cases are not consistently lower compared to the native cases.

## 4 Discussion

In this study, we apply the PSCOPE framework to conduct the implantation of a physical HeartMate 3 LVAD into a digital twin, obtaining a predictive model of the immediate post-implant hemodynamics. The observed validation residuals of these PSCOPE models demonstrate the potential of using this hybrid modeling method to aid clinicians in optimizing LVAD management for patients. A critical aspect of LVAD procedure planning involves anticipating the postoperative impact on the right heart, as post-LVAD RVF significantly increases morbidity and mortality rates of LVAD patients. The incidence of post-LVAD RVF, ranging from 5% to 44% depending on diagnostic criteria [39–42], highlights the need for thorough preoperative risk assessments. This study introduces a predictive modeling approach that allows clinicians to anticipate various hemodynamic indicators of post-LVAD right heart function (Appendix Fig. A1) for preoperative risk stratification of post-LVAD RVF.

The lack of a significant difference between native and adapted digital twin predictions (Tables 2 and 3) suggests that the changes in HR, SVR, and PVR do not substantially account for the discrepancies observed between predicted and clinical hemodynamics. This highlights the need for further investigation into additional physiological factors that could influence the accuracy of PSCOPE predictions. Such factors may include perioperative changes in other parameters related to cardiac function and vascular impedance. By identifying and understanding these influential factors, clinicians and researchers can better determine the contexts where PSCOPE is most effective for LVAD procedure planning, enhancing its clinical translational potential.

The accuracy of the predicted hemodynamics is also impacted by the protocol used to characterize the perioperative hemodynamic profile of the patients. Each clinically monitored hemodynamic parameter requires the consolidation of potentially thousands of measurements into a single value to characterize pre- and post-operative hemodynamics. The variability of each parameter is quantified by the standard deviation of its measurements, which can reflect both measurement noises and hemodynamic fluctuations over time. To minimize bias from transient physiological changes induced by perioperative treatments, it is ideal to focus on measurements obtained within the 12-hour windows immediately before and after LVAD implantation.

Moreover, the sampling frequency of the clinical measurements influences the extent to which the hemodynamics of the patient is sufficiently represented in the acquired data. Characteristic hemodynamic values derived from under-sampled datasets are prone to potential skewness in representing the underlying physiology, adversely impacting the accuracy of the derived PSCOPE predictions.

To enhance the personalization of a digital twin model, it is crucial to constrain the Lumped Parameter Network (LPN) with input values that accurately represent the patient’s physiology. These input values can be either directly calculated or systematically tuned based on the available patient data. In this study, HR, SVR, and PVR were the only LPN inputs directly calculated from the available clinical data, while other inputs were adjusted via iterative tuning. Systematic tuning enables personalization according to the completeness of the hemodynamic targets specified in the patient data. Thus, the extent of personalization achieved in the digital twin is closely related to the availability of comprehensive clinical data that includes all physiologically relevant hemodynamic parameters. Expanding the set of clinical measurements to capture additional hemodynamic parameters could improve the fidelity of the tuned LPN inputs, thereby enhancing personalization of the digital twin and potentially increasing the accuracy of predictions made by the PSCOPE framework.

Finally, it’s important to note that the time-varying elastance model of ventricular function utilized in the digital twins has limitations in accurately representing the interactions between the LVAD and the left ventricle (LV) [43–44]. Following implantation, the mechanics of the LV adapt to the loading conditions imposed by the operation of the LVAD, deviating from its native function. Therefore, future research should focus on developing a simulation that more precisely models the dynamics between the LVAD and the LV to enhance the PSCOPE framework’s capacity to predict post-LVAD hemodynamics.

## 5 Conclusion

In this study, we validated the PSCOPE framework by directly comparing its predictions against clinical data in three patient-specific cases of HeartMate 3 LVAD implantation. Our findings demonstrate PSCOPE’s potential to offer valuable insights into patient-specific physiology, enhancing preoperative risk stratification and surgical decision-making. We quantified PSCOPE’s ability to predict post-implant parameters such as CI, CVP, Pao, PAP, and PAWP. Despite encountering some challenges in predicting hemodynamic pulsatility accurately, the framework’s overall performance in replicating clinically observed trends underscores its potential clinical utility. We identified the LPN input parameter values and clinical data handling as potentially significant sources of discrepancy between predicted and clinical hemodynamics. This emphasizes the importance of comprehensive and precise perioperative physiology monitoring to acquire accurate hemodynamic targets, enabling accurate personalization of the digital twin within the PSCOPE framework.

In conclusion, the validation of the PSCOPE framework in patient-specific LVAD cases represents a significant step toward its integration into clinical practice. By providing clinicians with reliable predictions of post-surgical hemodynamics, treatment protocol optimization can improve LVAD patient outcomes. Continued research and refinement of this hybrid modeling approach holds promise for advancing personalized precision care in cardiovascular medicine.

## Data Availability

All data produced in the present study are available upon reasonable request to the authors

## A. Appendix

### Implementation of the lumped parameter network to generate digital twins

The LPN used in this study is a modified version of the Fontan circulation LPN developed by Kung et al. [45]. We expanded the cardiac model of the Fontan LPN to accommodate a bi-ventricular circulation and broadened its upper body simulation into parallel circuits for the head and hands. The equations describing the bi-ventricular LPN are reported below.

#### Atria

The pressure-volume relationship in the atria is described using active-passive equations that model the contraction, relaxation, and compliance of the chambers. The active pressure is scaled via an activation function (AA(t)) and added to the passive pressure as shown below:

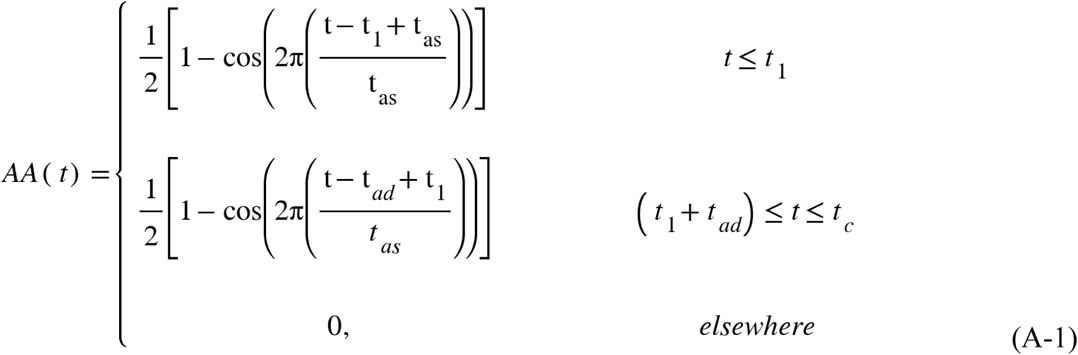

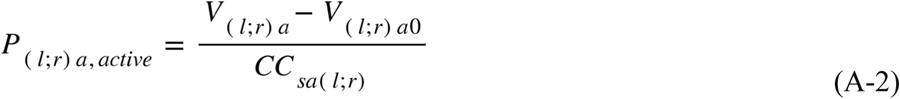

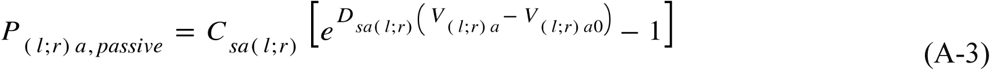

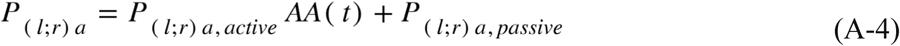

Where P_(l;r)a_ refers to pressure in the atrium; t_as_ is the atrial systolic time; t_c_ = 60/HR is the cardiac cycle time with HR values reported in Tables A1-A3; t_ad_ = t_c_ - t_as_ is the atrial diastolic time; t_1_ = 0.24*t_as_; t refers to a time point in the cardiac cycle; subscript “(l;r)” indicates the left or right atrium; CC_sa(l;r)_, C_sa(l;r)_, and D_sa(l;r)_, are constant parameters that describe the active-passive behavior of the atrium; V_(l;r)a_ and V_(l;r)a0_ represent atrial blood volume and reference volume, respectively.

#### Ventricles

The pressure-volume relationship in the ventricles is described using time-varying elastance equations as shown below:

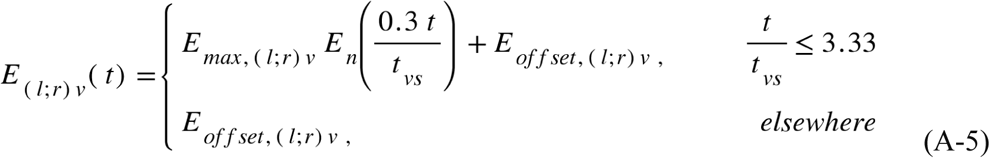

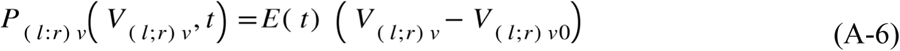

Where: P_(l;r)v_ refers to pressure in the ventricle; E_(l;r)v_(t) and E_n_ is the de-normalized and normalized elastance function, respectively; E_max,(l;r)v_,and E_offset,(l;r)v_ are constant parameters reported in Tables A1-A3; t_vs_ and t refer to ventricular systolic time, and a specific time point in the cardiac cycle, respectively; V_(l;r)v_ and V_(l;r)v0_ represent ventricular blood volume and reference volume, respectively; subscript “(l;r)” indicates the left or right ventricle.

#### Valves

The blood flow rate through the mitral, tricuspid, pulmonary, and aortic valves are described by the following equations:

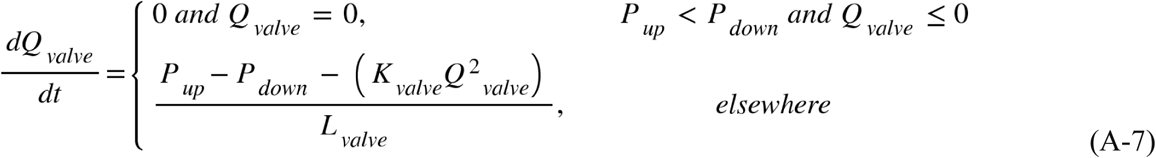

Where: Q_valve_ refers to transvalvular flow rate; P_up_ and P_down_ refer to upstream and downstream pressure of the valve; K_valve_ and L_valve_ are parameters characterizing valvular resistance and inductance, respectively.

#### Systemic circulation

The systemic circulation is modeled using resistors, capacitors, and inductors to represent viscous vascular resistance, vascular compliance, and blood flow momentum, respectively. The simulated hemodynamics is determined by the following equations that govern flow through these elements:

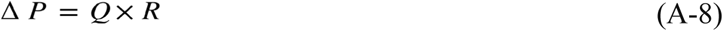

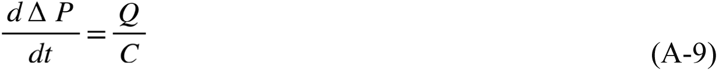

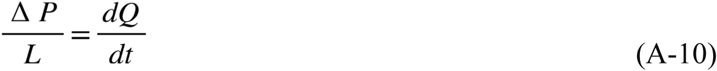

Where R, C, L, Δ*P*, and Q are the resistance value, capacitance value, inductance value, pressure drop across the element, and flow through the element, respectively.

**Table A1.** Parameter values for the native and adapted digital twins (Fig. 1A) generated to simulate the characterized preoperative hemodynamics of patient 1.

| Parameter | Native | Adapted | Native and Adapted |  |
| --- | --- | --- | --- | --- |
| HR (bpm) | 75 | 80 | Cardiac |  |
| Time Period (s) | | | $K_{tri} \text{ (mmHg.s}^2\text{.ml}^{-2}\text{)}$ | $5 \times 10^{-5}$ |
| $t_{vs}$ | 0.280 | 0.263 | $K_{pul} \text{ (mmHg.s}^2\text{.ml}^{-2}\text{)}$ | $3.5 \times 10^{-7}$ |
| $t_{as}$ | 0.240 | 0.225 | $K_{mit} \text{ (mmHg.s}^2\text{.ml}^{-2}\text{)}$ | $5 \times 10^{-5}$ |
| $t_l$ | 0.0576 | 0.0540 | $K_{ao} \text{ (mmHg.s}^2\text{.ml}^{-2}\text{)}$ | $2.5 \times 10^{-6}$ |
| Resistance (mmHg.s.ml<sup>-1</sup>) | | | $V_{rao} \text{ (ml)}$ | 10.0 |
| $R_{cora}$ | 3.4311 | 3.68505 | $V_{lao} \text{ (ml)}$ | 5.0 |
| $R_{corv}$ | 30.8799 | 33.16545 | $CC_{sar} \text{ (ml.mmHg}^{-1}\text{)}$ | 6.50 |
| $R_{crba}$ | 1.71031 | 1.83690 | $CC_{sal} \text{ (ml.mmHg}^{-1}\text{)}$ | 0.65 |
| $R_{crbv}$ | 6.06382 | 6.51263 | $C_{sar} \text{ (mmHg)}$ | 2.70 |
| $R_{ha}$ | 1.71031 | 1.83690 | $C_{sal} \text{ (mmHg)}$ | 0.270 |
| $R_{hv}$ | 6.06382 | 6.51263 | $D_{sar} \text{ (ml}^{-1}\text{)}$ | 0.03375 |
| $R_{svc}$ | 0.07898 | 0.08482 | $D_{sal} \text{ (ml}^{-1}\text{)}$ | 0.225 |
| $R_{thao}$ | 0.12340 | 0.13253 | $E_{max,rv} \text{ (mm.Hg.ml}^{-1}\text{)}$ | 0.08808 |
| $R_{abao}$ | 0.12340 | 0.13253 | $E_{max,lv} \text{ (mm.Hg.ml}^{-1}\text{)}$ | 0.88074 |
| $R_{lega}$ | 1.39243 | 1.49549 | $E_{offset,rv} \text{ (mm.Hg.ml}^{-1}\text{)}$ | 0.01922 |
| $R_{legc}$ | 3.21331 | 3.45114 | $E_{offset,lv} \text{ (mm.Hg.ml}^{-1}\text{)}$ | 0.19430 |
| $R_{legv}$ | 0.74977 | 0.80527 | $V_{rvo} \text{ (ml)}$ | 6.96094 |
| $R_{abivc}$ | 0.01234 | 0.01325 | $V_{lvo} \text{ (ml)}$ | -23.49266 |
| $R_{thivc}$ | 0.01234 | 0.01325 | Inductance (mmHg.s<sup>2</sup>.ml<sup>-1</sup>) X 10<sup>-6</sup> | |
| $R_{la}$ | 1.86579 | 2.00389 | $L_{tri}$ | 1.0 |
| $R_{lv}$ | 2.79869 | 3.00583 | $L_{pul}$ | 5.0 |
| $R_{ka}$ | 1.97438 | 2.12051 | $L_{mit}$ | 1.0 |
| R<sub>kv</sub> | 2.96157 | 3.18077 | L<sub>ao</sub> | 5.0 |
| R<sub>ia</sub> | 2.51734 | 2.70366 | L<sub>crba</sub> | 120.0 |
| R<sub>iv</sub> | 2.51734 | 2.70366 | L<sub>ha</sub> | 120.0 |
| R<sub>lpa</sub> | 0.10199 | 0.04443 | L<sub>thao</sub> | 200.0 |
| R<sub>rpa</sub> | 0.10199 | 0.04443 | L<sub>abao</sub> | 100.0 |
| R<sub>lpv</sub> | 0.42105 | 0.18342 | L<sub>lega</sub> | 200.0 |
| R<sub>rpv</sub> | 0.42105 | 0.18342 | **Capacitance (ml.mmHg<sup>-1</sup>)** |  |
| R<sub>aov</sub> | 0.01 | 0.01 | C<sub>cor</sub> | 0.00063 |
| **Capacitance (ml.mmHg<sup>-1</sup>) cont'd** |  |  | C<sub>ccrb</sub> | 2.11723 |
| C<sub>abivc</sub> | 3.24802 |  | C<sub>h</sub> | 1.92475 |
| C<sub>thivc</sub> | 6.49604 |  | C<sub>svc</sub> | 5.41337 |
| C<sub>l</sub> | 3.60891 |  | C<sub>ao</sub> | 0.35196 |
| C<sub>k</sub> | 3.60891 |  | C<sub>thao</sub> | 0.19248 |
| C<sub>i</sub> | 4.33070 |  | C<sub>abao</sub> | 0.09624 |
| C<sub>la</sub> | 0.68436 |  | C<sub>lega</sub> | 0.630 |
| C<sub>ra</sub> | 0.68436 |  | C<sub>legv</sub> | 2.36250 |

**Table A2.**
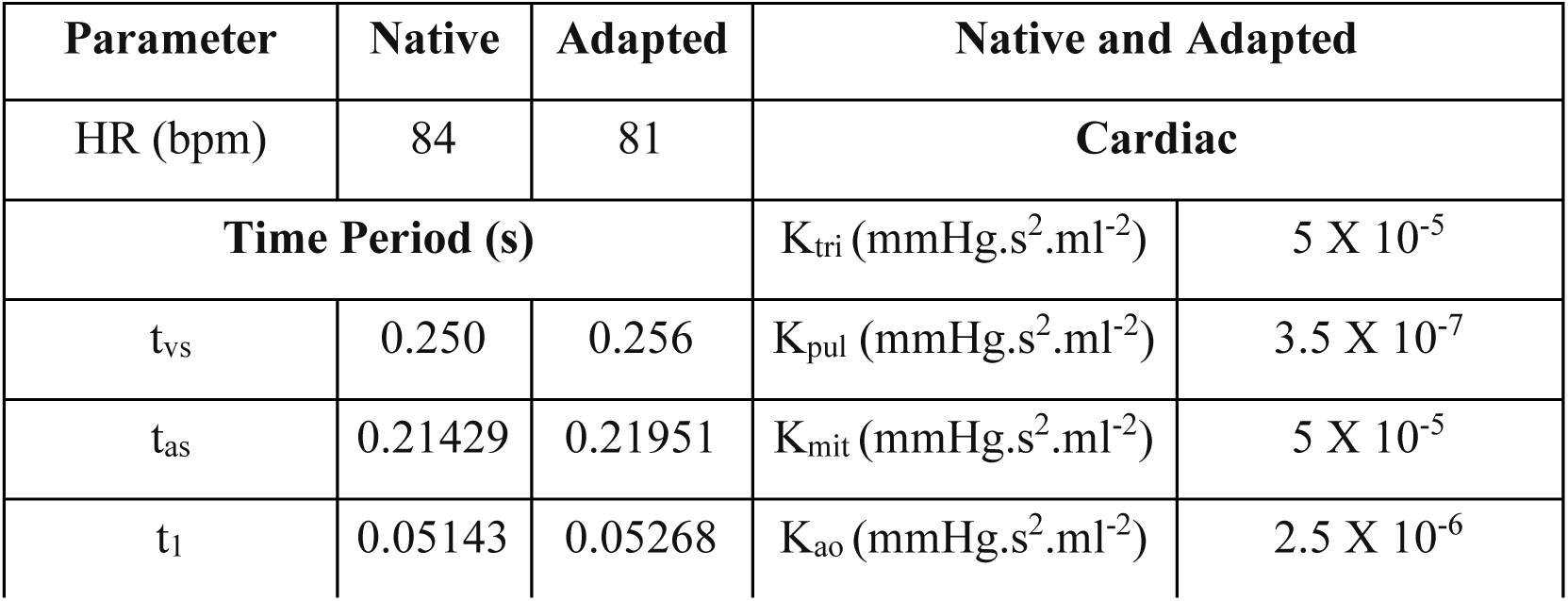
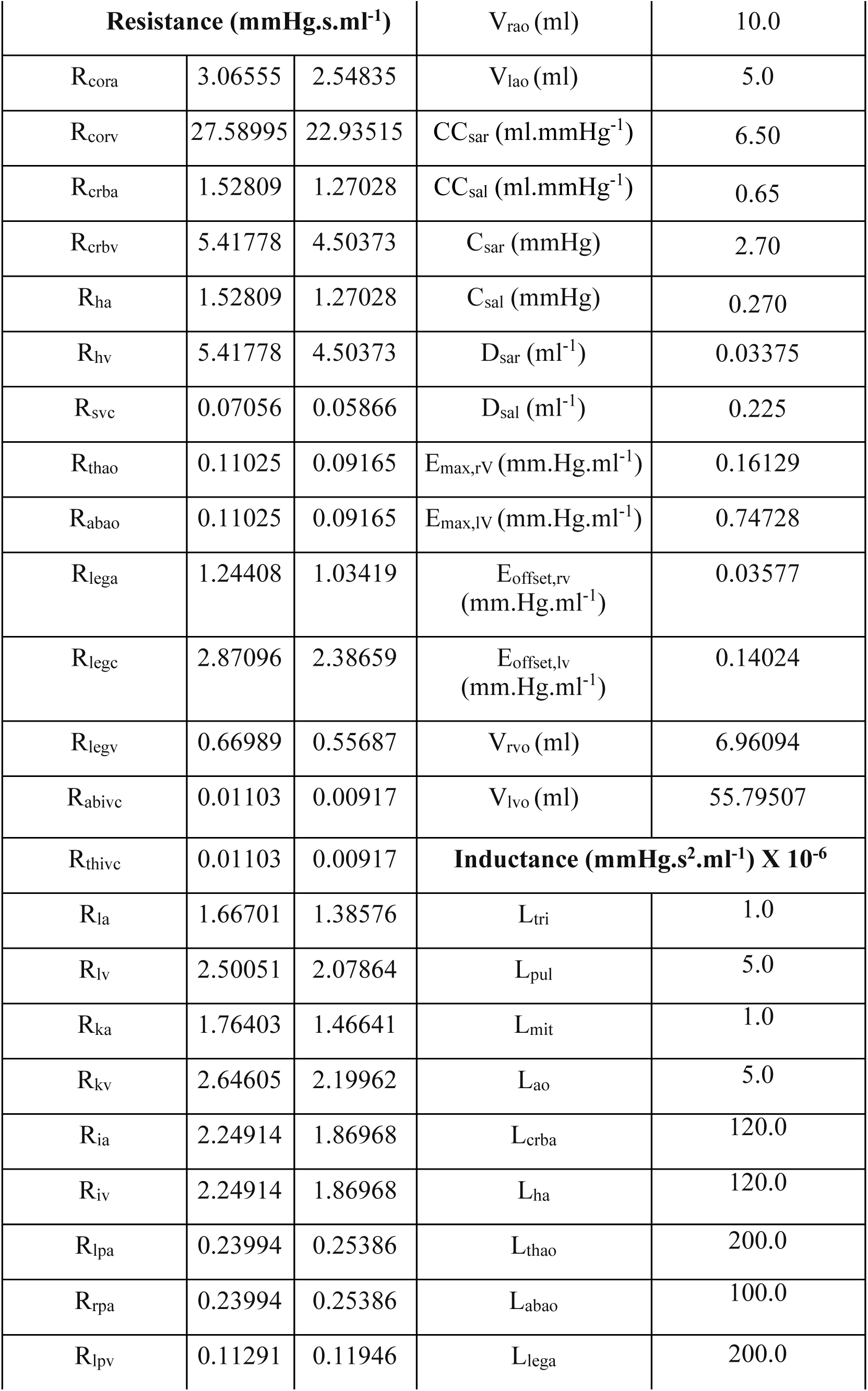
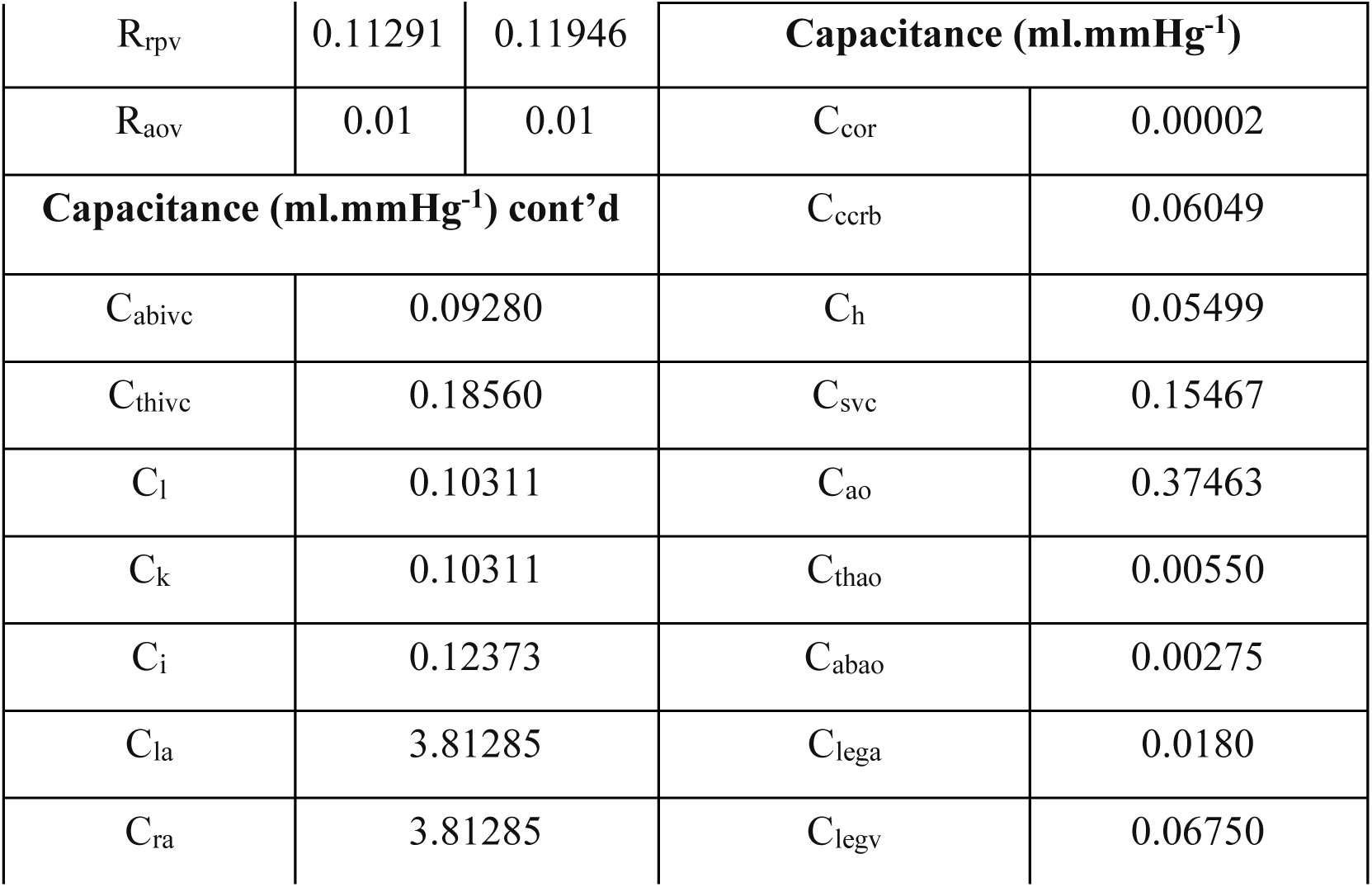
LPN parameter values for the native and adapted digital twins (Fig. 1A) generated to simulate the characterized preoperative hemodynamics of patient 2.

**Table A3.** LPN parameter values for the native digital twin (Fig. 1A) generated to simulate the characterized preoperative hemodynamics of patient 3. The adapted digital twin could not be generated due to insufficient clinical data.

| Parameter | Native |  |  |
| --- | --- | --- | --- |
| Time Period (s) |  | Cardiac |  |
| $t_{vs}$ | 0.22581 | HR (bpm) | 93 |
| $t_{as}$ | 0.19355 | $K_{tri}$ (mmHg.s<sup>2</sup>.ml<sup>-2</sup>) | $5 \times 10^{-5}$ |
| $t_l$ | 0.04645 | $K_{pul}$ (mmHg.s<sup>2</sup>.ml<sup>-2</sup>) | $3.5 \times 10^{-7}$ |
| **Resistance (mmHg.s.ml<sup>-1</sup>)** | | $K_{mit}$ (mmHg.s<sup>2</sup>.ml<sup>-2</sup>) | $5 \times 10^{-5}$ |
| $R_{cora}$ | 2.370 | $K_{ao}$ (mmHg.s<sup>2</sup>.ml<sup>-2</sup>) | $2.5 \times 10^{-6}$ |
| $R_{corv}$ | 21.330 | $V_{rao}$(ml) | 10.0 |
| $R_{crba}$ | 1.18138 | $V_{lao}$(ml) | 5.0 |
| $R_{crbv}$ | 4.18853 | $CC_{sar}$ (ml.mmHg<sup>-1</sup>) | 6.50 |
| $R_{ha}$ | 1.18138 | $CC_{sal}$ (ml.mmHg<sup>-1</sup>) | 0.65 |
| $R_{hv}$ | 4.18853 | $C_{sar}$ (mmHg) | 2.70 |
| $R_{svc}$ | 0.05455 | $C_{sal}$ (mmHg) | 0.270 |
| $R_{thao}$ | 0.08524 | $D_{sar}$ (ml<sup>-1</sup>) | 0.03375 |
| $R_{abao}$ | 0.08524 | $D_{sal}$ (ml<sup>-1</sup>) | 0.225 |
| $R_{lega}$ | 0.96181 | $E_{max,rv}$ (mm.Hg.ml<sup>-1</sup>) | 0.31977 |
| $R_{legc}$ | 2.21956 | $E_{max,lv}$ (mm.Hg.ml<sup>-1</sup>) | 0.27254 |
| $R_{legv}$ | 0.51790 | $E_{offset,rv}$<br>(mm.Hg.ml<sup>-1</sup>) | 0.02839 |
| $R_{abivc}$ | 0.00852 | $E_{offset,lv}$<br>(mm.Hg.ml<sup>-1</sup>) | 0.04798 |
| $R_{thivc}$ | 0.00852 | $V_{rvo}$ (ml) | 6.96094 |
| $R_{la}$ | 1.28878 | $V_{lvo}$ (ml) | 39.15444 |
| $R_{lv}$ | 1.93317 | **Inductance (mmHg.s<sup>2</sup>.ml<sup>-1</sup>) X 10<sup>-6</sup>** | |
| $R_{ka}$ | 1.36379 | $L_{tri}$ | 1.0 |
| $R_{kv}$ | 2.04568 | $L_{pul}$ | 5.0 |
| $R_{ia}$ | 1.73883 | $L_{mit}$ | 1.0 |
| $R_{iv}$ | 1.73883 | $L_{ao}$ | 5.0 |
| $R_{lpa}$ | 0.12852 | $L_{crba}$ | 120.0 |
| $R_{rpa}$ | 0.12852 | $L_{ha}$ | 120.0 |
| $R_{lpv}$ | 0.13112 | $L_{thao}$ | 200.0 |
| $R_{rpv}$ | 0.13112 | $L_{abao}$ | 100.0 |
| $R_{aov}$ | 0.01 | $L_{lega}$ | 200.0 |
| **Capacitance (ml.mmHg<sup>-1</sup>)** |  |  |  |
| $C_{thivc}$ | 43.30698 | $C_{cor}$ | 0.00420 |
| $C_l$ | 24.05944 | $C_{ccrb}$ | 2.11723 |
| $C_k$ | 24.05944 | $C_h$ | 1.92475 |
| $C_i$ | 28.87132 | $C_{svc}$ | 5.41337 |
| $C_{la}$ | 2.72346 | $C_{ao}$ | 0.69326 |
| $C_{ra}$ | 2.72346 | $C_{thao}$ | 1.28317 |
| $C_{abivc}$ | 21.65349 | $C_{abao}$ | 0.64158 |
| $C_{legv}$ | 15.750 | $C_{lega}$ | 4.20 |

**Fig. A1.**
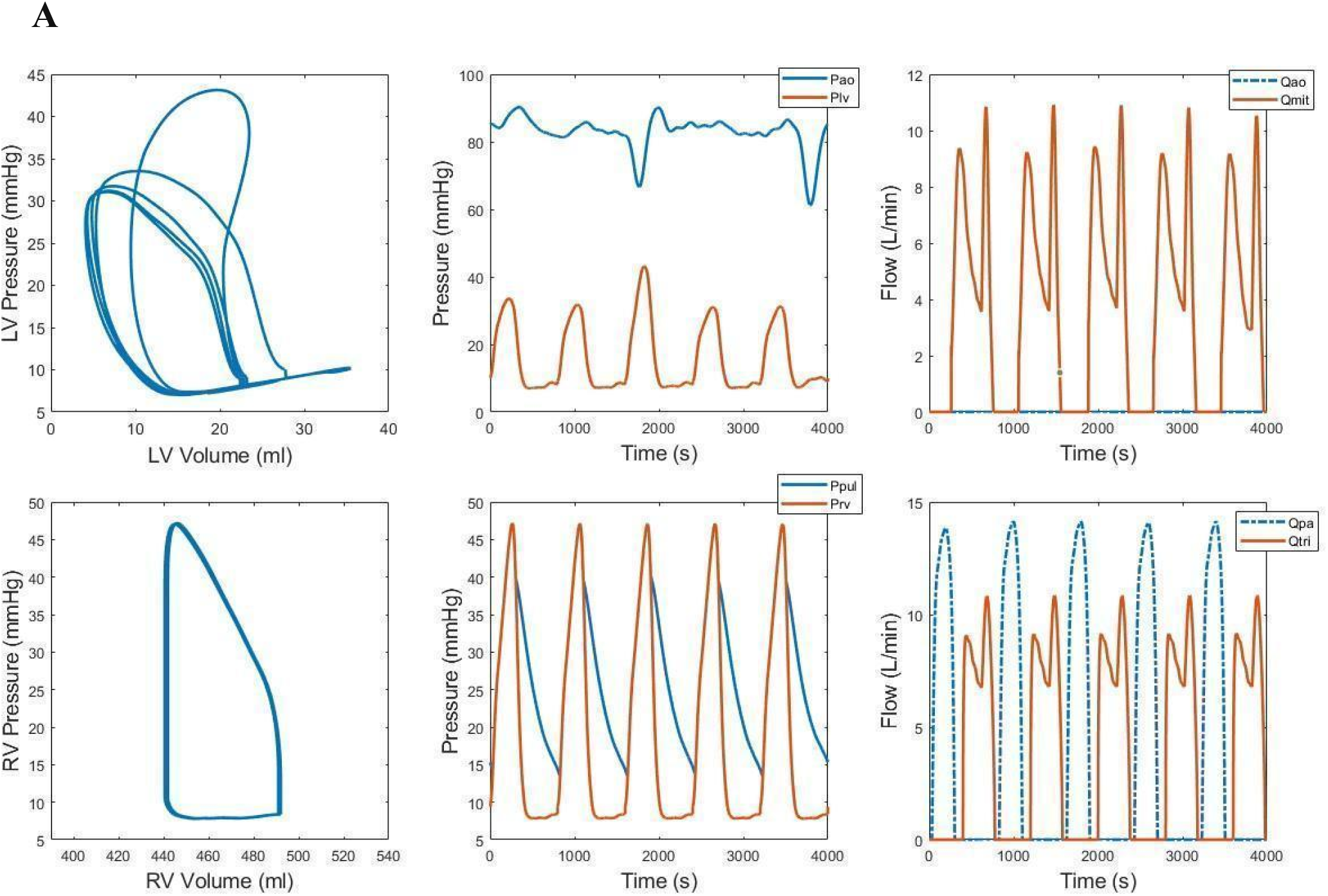
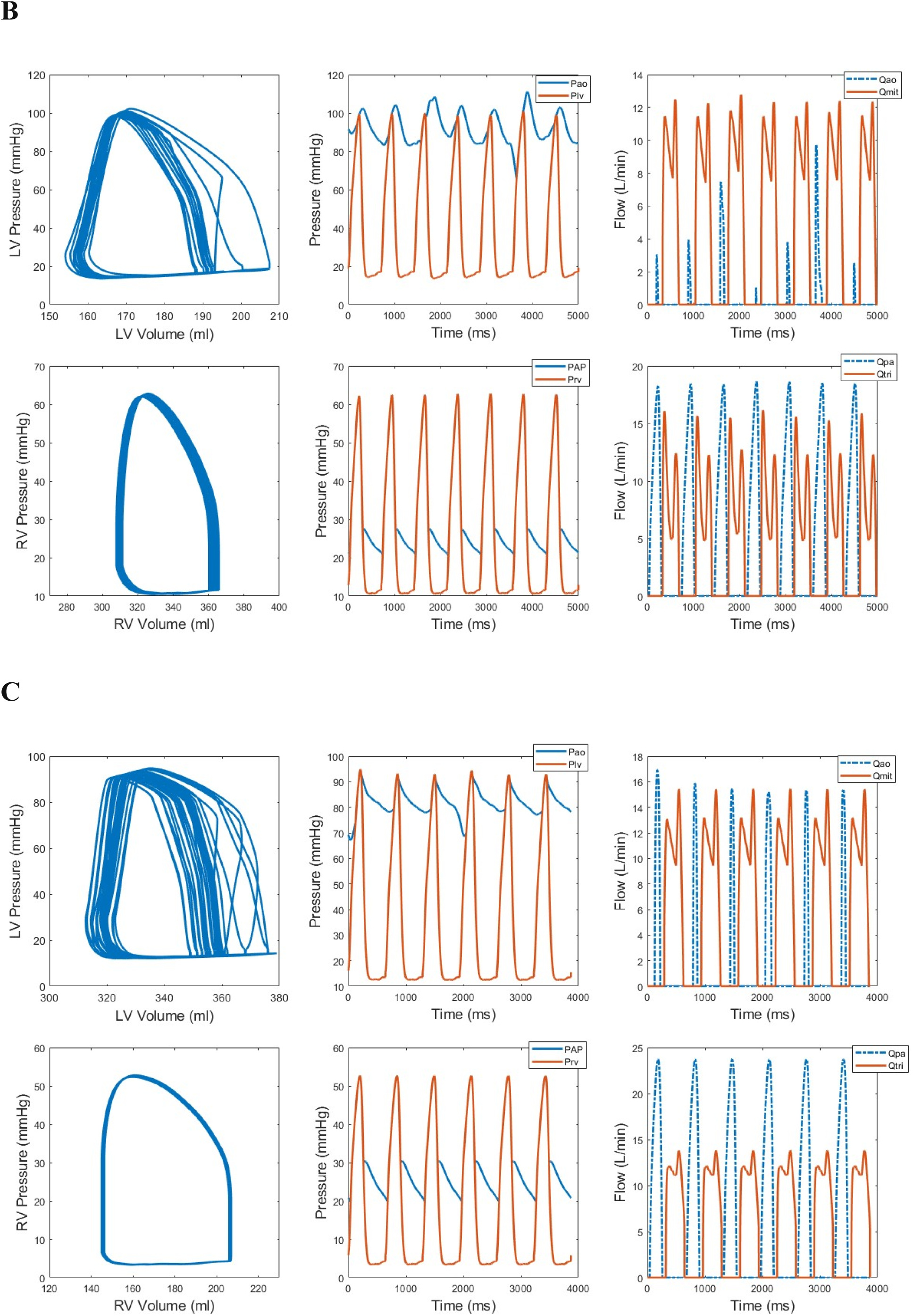
PSCOPE predictions of postoperative hemodynamics in the left and right heart for patient 1 (A), patient 2 (B), and patient 3 (C). LV-Left Ventricle; RV- Right Ventricle; Pao-Aortic pressure; Plv- Left ventricular pressure; Qao- Transaortic flow rate; Qmit- Transmitral flow rate; PAP- Pulmonary arterial pressure; Prv- Right ventricular pressure; Qpa- Flow rate through the pulmonary valve; Qtri-Flow rate through the tricuspid valve.

## Acknowledgements

We acknowledge Abbott’s generous provision of a HeartMate3 device for this study.

## Author Contributions

**Abraham Umo**: Methodology, Software, Validation, Formal analysis, Investigation, Data curation, Visualization, Writing-original draft preparation, Writing-review and editing, Project administration **Brett Welch**: Resources, Data curation **Arman Kilic**: Conceptualization, Resources **Ethan Kung**: Conceptualization, Methodology, Software, Validation, Formal Analysis, Resources, Supervision, Funding Acquisition, Visualization, Writing-review and editing, Project administration.

